# Interim Analysis of Pandemic Coronavirus Disease 2019 (COVID-19) and the SARS-CoV-2 virus in Latin America and the Caribbean: Morbidity, Mortality and Molecular Testing Trends in the Region

**DOI:** 10.1101/2020.04.25.20079863

**Authors:** Katherine Simbaña-Rivera, Lenin Gómez-Barreno, Jhon Guerrero, Fernanda Simbaña-Guaycha, Raúl Fernández, Andrés López-Cortés, Alex Lister, Esteban Ortiz-Prado

**Affiliations:** One Health Research Group, Faculty of Medicine, Universidad de Las Américas, Quito, Ecuador; Scientific Association of Medical Students, Universidad Central del Ecuador, Quito, Ecuador; Centro de Investigación Genética y Genómica, Facultad de Ciencias de la Salud Eugenio Espejo, Universidad UTE, Quito 170129, Ecuador; Red Latinoamericana de Implementación y Validación de Guías Clínicas Farmacogenómicas (RELIVAF-CYTED), Quito, Ecuador; Faculty of Medicine, University of Southampton, Southampton, United Kingdom

**Keywords:** COVID-19, Coronavirus, Latin America, Epidemiology, Case fatality rate

## Abstract

**Background:** The relentless advance of the SARS-CoV-2 virus pandemic has resulted in a significant burden on countries, regardless of their socio-economic conditions. The virus has infected more than 2.5 million people worldwide, causing to date more than 150,000 deaths in over 210 countries.

**Objective:** The aim of this study was to describe the trends in cases, tests and deaths related to novel coronavirus disease (COVID-19) in Latin American and Caribbean (LAC) countries.

**Methodology:** Data were retrieved from the WHO-Coronavirus Disease (COVID-2019) situation reports and the Center for Systems Science and Engineering (CSSE) databases from Johns Hopkins University. Descriptive statistics including death rates, cumulative mortality and incidence rates, as well as testing rates per population at risk were performed. A comparison analysis among countries with ≥50 confirmed cases was performed from February 26^th^, 2020 to April 8^th^, 2020.

**Results:** Brazil had the greatest number of cases and deaths in the region. Panama experienced a rapid increase in the number of confirmed cases with Trinidad and Tobago, Bolivia and Honduras having the highest case fatality rates. Panama and Chile conducted more tests per million inhabitants and more tests per day per million inhabitants, followed by Uruguay and El Salvador. Dominican Republic, Bolivia, Ecuador and Brazil had the highest positive test rates.

**Conclusions:** The COVID-19 disease pandemic caused by the SARS-CoV-2 virus has progressed rapidly in LAC countries. Some countries have been affected more severely than others, with some adopting similar disease control methods to help slow down the spread of the virus. With limited testing and other resources, social distancing is needed to help alleviate the strain on already stretched health systems.

## Background

Communicable diseases have long been part of our humanity, having an impact on global population levels, human culture and socialization [1]. Bacterial and viral infections are responsible for millions of deaths worldwide, with pandemics over history including the black death, smallpox, cholera, AIDS and influenza-type disease. Recent respiratory disease outbreaks include the Severe Acute Respiratory Syndrome (SARS), the Middle-East Respiratory Syndrome (MERS) and the latest, the 2019 novel Coronavirus disease (termed COVID-19) caused by the SARS-CoV-2 virus [2]. The current classification of coronaviruses recognizes 39 species in 27 subgenera that belong to the family *Coronaviridae*, of which three can cause disease in both animals and humans [2, 3]. SARS-CoV-2 is an enveloped virus with roughly spherical virions of approximately 60-140nm in diameter [4]. The first genome of SARS-CoV-2 named Wuhan-Hu-1 (NCBI reference sequence NC_045512) was isolated and sequenced in China [4, 5].

On December 31^st^ 2019, a number of atypical pneumonia-like illness cases were reported in Hubei province, China [6]. On January 10^th^, through genetic sequencing, it was confirmed that this pneumonia was caused by a novel coronavirus (SARS-CoV-2). Ten days later, the first confirmed cases outside mainland China occurred in Japan, South Korea and Thailand. By the end of January, at least 17 people had died and more than 570 others had been infected worldwide [6, 7]. After only three months, on March 11^th^, the staggering number of cases led the World Health Organization (WHO) declaring the outbreak a pandemic [8] and since then, its distribution has been relentless, infecting more than 2.2 million people and killing more than 150,000 people worldwide [9].

The virus spread first in Asia, then to Europe and North America and finally made its way to South America and Africa. The impact of the atypical respiratory disease has already put substantial pressure on health systems globally [10–12]. This pressure is due to the percentage of patients who require hospitalization and the speed at which symptoms can worsen [6]. For Latin America, early cases were able to be managed, however as the number of cases rose, countries with little investment into public health struggled to control the number of cases. The first country to confirm a COVID-19 case in Latin America was Brazil, followed by Mexico and later Ecuador. All Latin American countries have reported the disease at the time of writing [9].

The aim of this study was to describe the trends of COVID-19 in each Latin American and the Caribbean (LAC) countries through the calculation of different rates that reflect the reality of each one. These were then compared with the rates seen in China and Italy recorded up until April, 8^th^ 2020.

## Material and Methods

### Study Design

A descriptive observational study using secondary international reports was carried out for all Latin American and Caribbean countries from February 26^th^, 2020 to April 8th, 2020 that showed more than 50 confirmed cases by April 8^th^. Furthermore, China for being the first country to report cases, and Italy for being the European country with highest mortality reported, were added to the analysis from January 21^st^, 2020 to April 8^th^, 2020 as comparison models.

### Source of Data

Secondary data was retrieved from the three main publicly available databases that are publishing information about COVID-19. We use the daily reports from WHO-Coronavirus Disease (COVID-19) situation reports [13], the Center for Systems Science and Engineering (CSSE) at Johns Hopkins University [14] and the on-line data repository https://ourworldindata.org/coronavirus-source-data [15].

Sequences and amino acid replacement of SARS-CoV-2 virus information were retrieved from the Global Initiative on Sharing All Influenza Data (GISAID) (https://www.gisaid.org/), an initiative that promotes the international sharing of SARS-CoV-2 virus sequences to better understand how the novel coronavirus evolves, spreads and how it causes pathology [16, 17] as well as the CoV-GLUE Project (http://cov-glue.cvr.gla.ac.uk/#/home). The CoV-GLUE Project is a database of amino acid replacements that have been observed in sequences sampled from the pandemic strain [18]. Lastly, Supplementary Table 1 details the knowledge of all contributions and laboratories of this study according to GISAID requirement.

### Study Size

All confirmed COVID-19 countries with more than fifty cases reported in the region up to April 8^th^ were included in the analysis.

### Bias

To reduce the risk of observer bias, data collection was performed using the triangulation technique with three researchers of our team. Using this technique described by Carvalho and White [19], it aided in the reduction of bias as any errors in data collection could be highlighted through comparison between analysts.

### Variables

The number of confirmed cases and deaths attributed to COVID-19 were transformed into crude and cumulative numbers to obtain cumulative confirmed cases (CCC) and cumulative confirmed deaths (CCD) per country.

The morbidity and mortality incidence, case-fatality rate, positive-test rate and test performed were calculated according to the entire population per country.

### Statistical Analysis

The rate of cumulative incidence of confirmed cases (CICC) and deaths (CICD) per million inhabitants was calculated to reflect the impact of the coronavirus for each country using the population projection for 2020, according to the World Population Prospects 2019 [20].

The case fatality rate (CFR%) and positive test rate (PTR%) with a confidence interval of 95% for crude rates were computed for each country by April 8^th^ as follow:

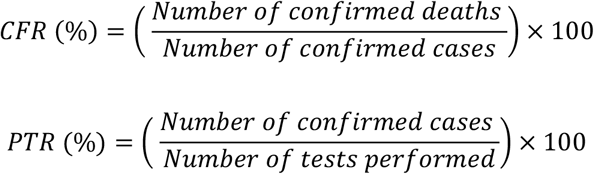

For an approach related to the test performed in each country. The influence of population size was reduced by the calculation of tests per million inhabitants (TMI) and to control time distribution daily tests per million inhabitants (DTMI) with the formulas:

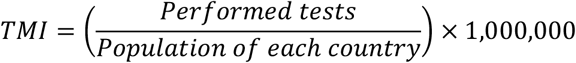

Additionally, to control the time distribution of cases per population and time since the first report, the DCMI was calculated:

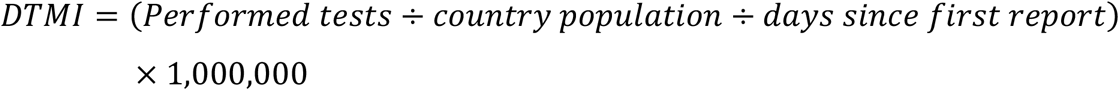

Finally, results were grouped as confirmed case-related data (CCC, CICC, DCMI), death-related data (CCD, CICD, CFR%) and test-related data (TMI, DTMI and PTR%).

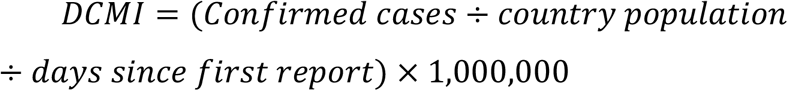

## Results

In the first 40 days since the first LAC countries’ report, the number of cases in the region reached 44,233 with more than 1,770 deaths. The total number of cases reported in the region until early-April 2020 represents 1% of the total number of confirmed cases worldwide.

### Confirmed Case-Related Data Analysis

#### Cumulative Incidence of Confirmed Cases (CCC)

Most countries in the region have a similar CCC trend compared to Italy. On April 8^th^, Brazil, Chile and Ecuador had the highest number of CCC with 16,170 cases, 5,546 cases and 4,450 cases respectively, which correspond to the 42^nd^, 36^th^ and 38^th^ days since their first report. At the same time, Honduras, Costa Rica and Venezuela had the lowest reports with 312 cases, 502 cases and 167 cases respectively, which correspond to the 25^th^, 27^th^ and 31^st^ days since their first report (Figure 1).

**Figure 1.**
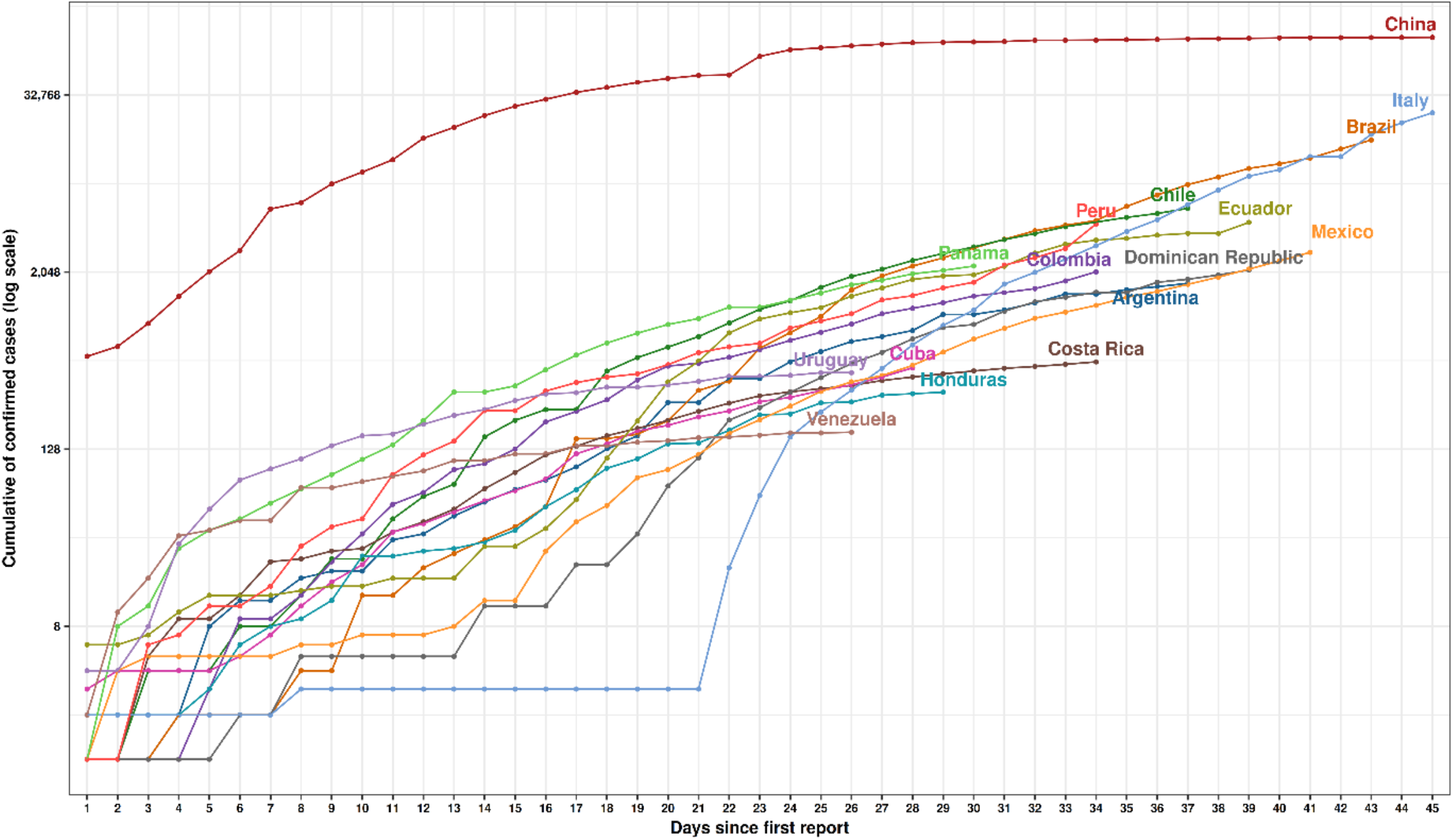
Cumulative of confirmed cases since the first case reported in each country of Latin America and the Caribbean.

This data was obtained from the daily new cases registered by country as shown in Figure 2, which demonstrated that most of them reached its highest value on April 8^th^. For example, Brazil highest peak was 2,136 cases, followed by Peru (1,388), Ecuador (703), Chile (430) and Mexico (346) (Figure 2).

**Figure 2.**
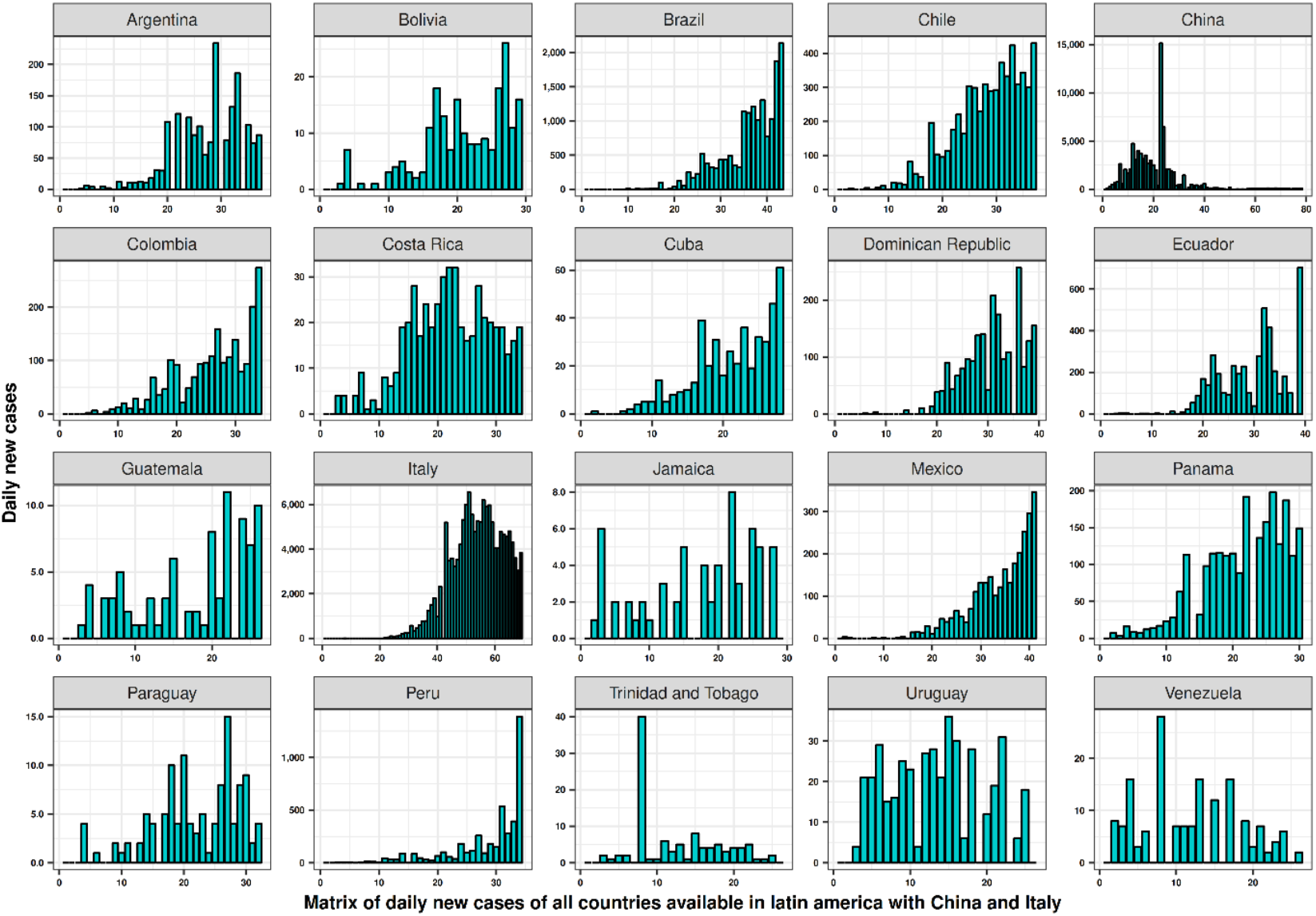
Matrix of daily new cases since the first case reported in each country of Latin America and the Caribbean. Italy and China are depicted in darker green to show as a comparison.

#### Cumulative Incidence of Confirmed Cases per Million Inhabitants (CICC)

The cumulative incidence of confirmed cases reached 10 per million people in Italy at the 37^th^ day (March 8^th^) since the first official report of COVID-19, while as a reference, China reported a plateau of 5 per million inhabitants since the 25^th^ day (February 16^th^). In this sense, some Latin American Countries like Panama, Chile and Ecuador show faster trends in confirmed cases compared to Italy and China (Figure 3).

**Figure 3.**
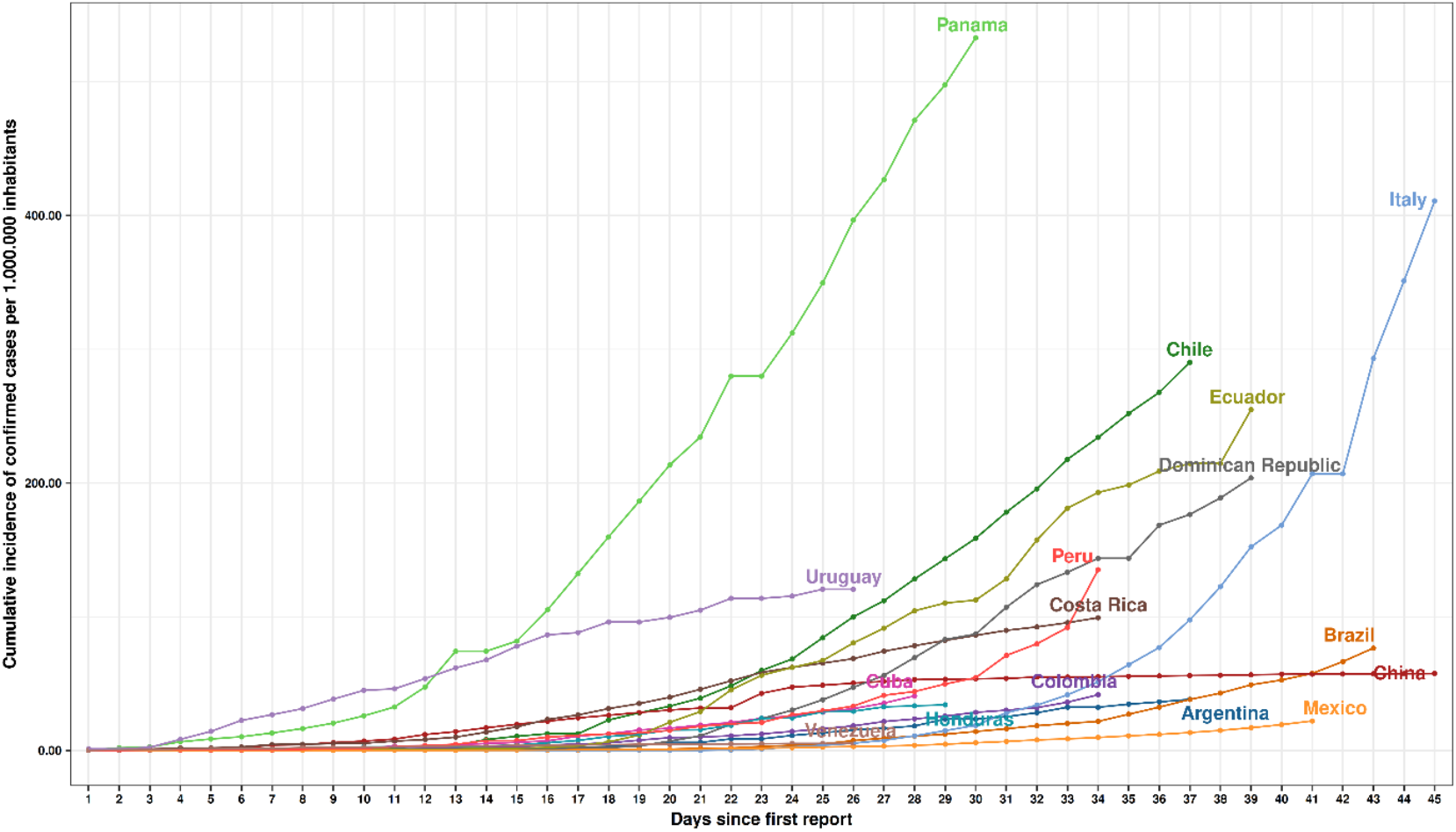
Cumulative incidence of confirmed cases per million inhabitants since the first case reported in each country of Latin America and the Caribbean.

Panama reached a rate of 10 per million as fast as the 15^th^ day (March 25^th^) since the first official report and in only 11 days, this number increased by 300% to a rate of 40 cases per million people. In the same way, Uruguay had a rapid increase in the incidence rate in only 20 days, reaching 10 cases per million people. Chile reported a slower start with a CICC of 10 cases per one million on the 26^th^ day (March 29^th^) after its first report, a value that got duplicated in only 6 days up to 20 cases per million on April 1^st^. Ecuador reached a CICC of 10 per million on the 27^th^ day (March 28^th^) of the outbreak, with this figure doubling 13 days later on April 5^th^.

Countries with higher population density had fewer rates including Brazil (6.2 per million), Mexico (2.1 per million) and Argentina (3.9 per million) inhabitants on the first 40 days after its first official case was reported in relationship with less populate countries.

#### Daily Confirmed Cases per Million Inhabitants (DCMI)

On April 8^th^, Italy reached 34 confirmed cases per million inhabitants (DCMI) daily, 68 days after its first official report. This number is still lower for Latin-American countries, such as Panama, that with only 29 days after the initial case had already reached 17.9 daily confirmed cases per million people, followed by Chile with 8 (36^th^ day), Ecuador 6.6 (38^th^ day), the Dominican Republic 5.1 (38^th^ day) and Uruguay 4.9 (25^th^ day) (Table 1).

**Table 1.**
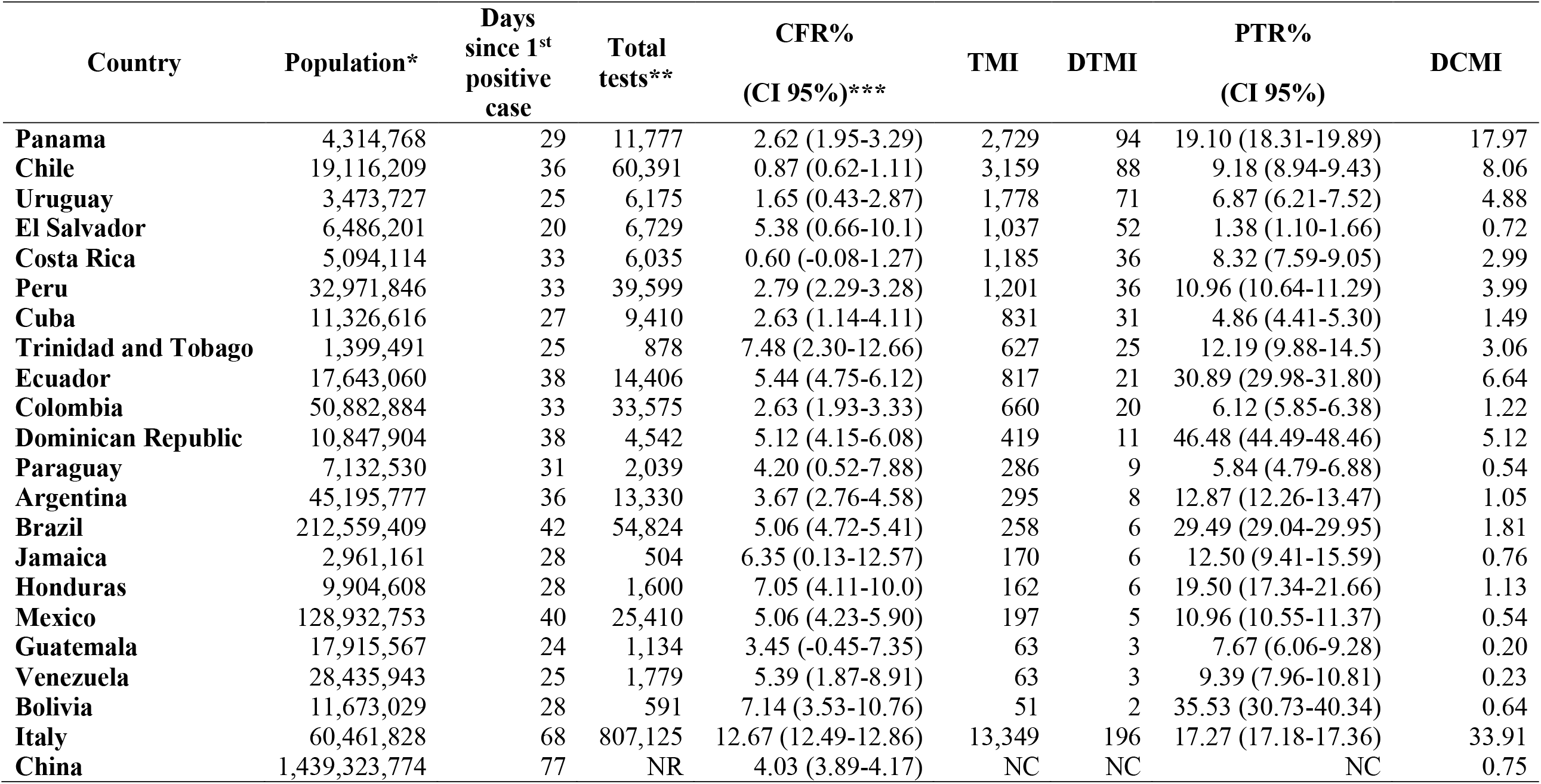
Comparison of CFR%, TMI, DTMI, PTR%, DCMI between China, Italy and the rest of Latin American and Caribbean countries with more than 50 confirmed cases until April 8th. CFR% = Case fatality rate; TMI = Tests per million inhabitants; DTMI = Daily test per million inhabitants; PTR% = Positive test rate; DCMI = Daily confirmed cases per million inhabitants; NR = Non reported; NC = Non calculated. Sources: *https://population.un.org/wpp/Download/Standard/Population; **https://ourworldindata.org/coronavirus-source-data; ***CFR calculated from Johns Hopkins University data base. + Days calculated until 08-04-2020

### Confirmed Death Case-Related Data Analysis

#### Cumulative Number of Confirmed Death (CCD)

The number of deaths reached an exponential growth, especially in Italy, reaching 1,809 CCD after 45 days since its first report. In comparison, cases reported by April 8^th^ in Brazil reaching 819 reports (40^th^ day), while Ecuador topped 242 (38^th^ day), Mexico 141 (40^th^ day), Peru 121 (33^rd^ day) and the Dominican Republic 108 (38^th^ day) (Figure 4).

**Figure 4.**
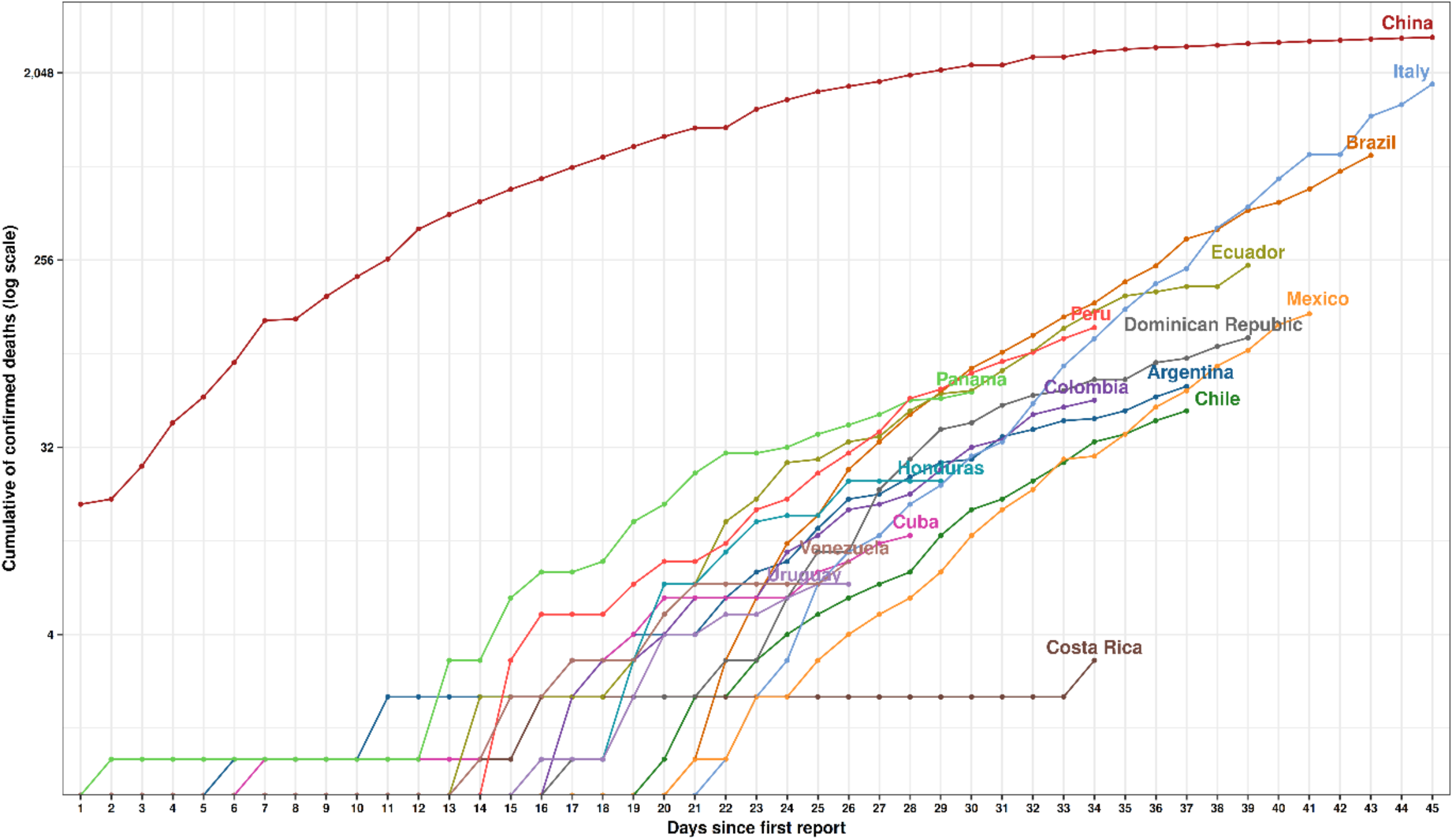
Cumulative of confirmed cases since the first death reported in each country of Latin America and the Caribbean.

Brazil had the highest peak of daily new deaths in the period, with 133 deaths by April 8^th^, followed by Ecuador with 51 deaths on the same day. Mexico reached the highest peak of deaths on April 7^th^ with 31, Peru on April 2^nd^ with 17 and the Dominican Republic peaked 14 deaths on April the 5^th^ (Figure 5).

**Figure 5.**
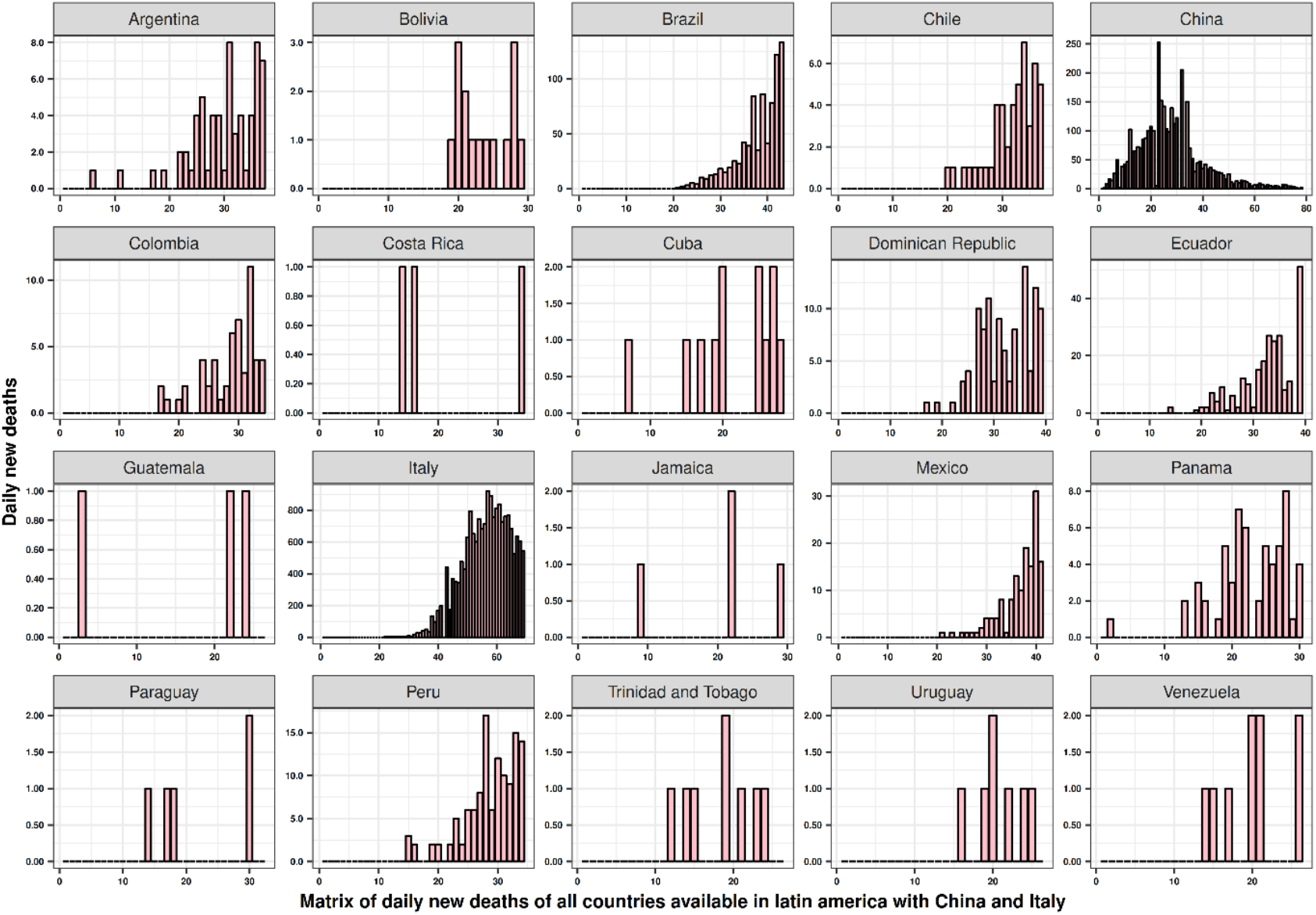
Matrix of daily new deaths since the first case reported in each country of Latin America and the Caribbean.

#### Cumulative incidence of deaths per million inhabitants (CICD)

In Italy, the cumulative incidence of confirmed deaths per million inhabitants reached 10 death per million people after 40 days after the first official report (March 11^th^). Only five days later, this number duplicated. In contrast, China showed a plateau since the 34^th^ day (February 25^th^) after the first case was reported, reaching less than 2.3 deaths per million inhabitants.

In Latin America, countries such as Panama and Ecuador reached 10 deaths per million inhabitants as fast as the 26^th^ (April 5^th^) and the 35^th^ day (April 5^th^) after the first case was reported, respectively. The Dominican Republic reached 9 cumulative incidence death rate per million inhabitants on the 38^th^ day (April 8^th^) after the first case report.

Other countries had a slower start, reaching a lower CICD rate after the 34th day of the initial report as Chile (2.2 per million), Argentina (1.3 per million), Mexico (1.1 per million) and Brazil (3.1 per million) on the 34^th^ (April 6^th^), 35^th^ (April 7^th^), 39^th^ (April 7^th^) and 41^st^ (April 7^th^) days after the initial report, respectively (Figure 6).

**Figure 6.**
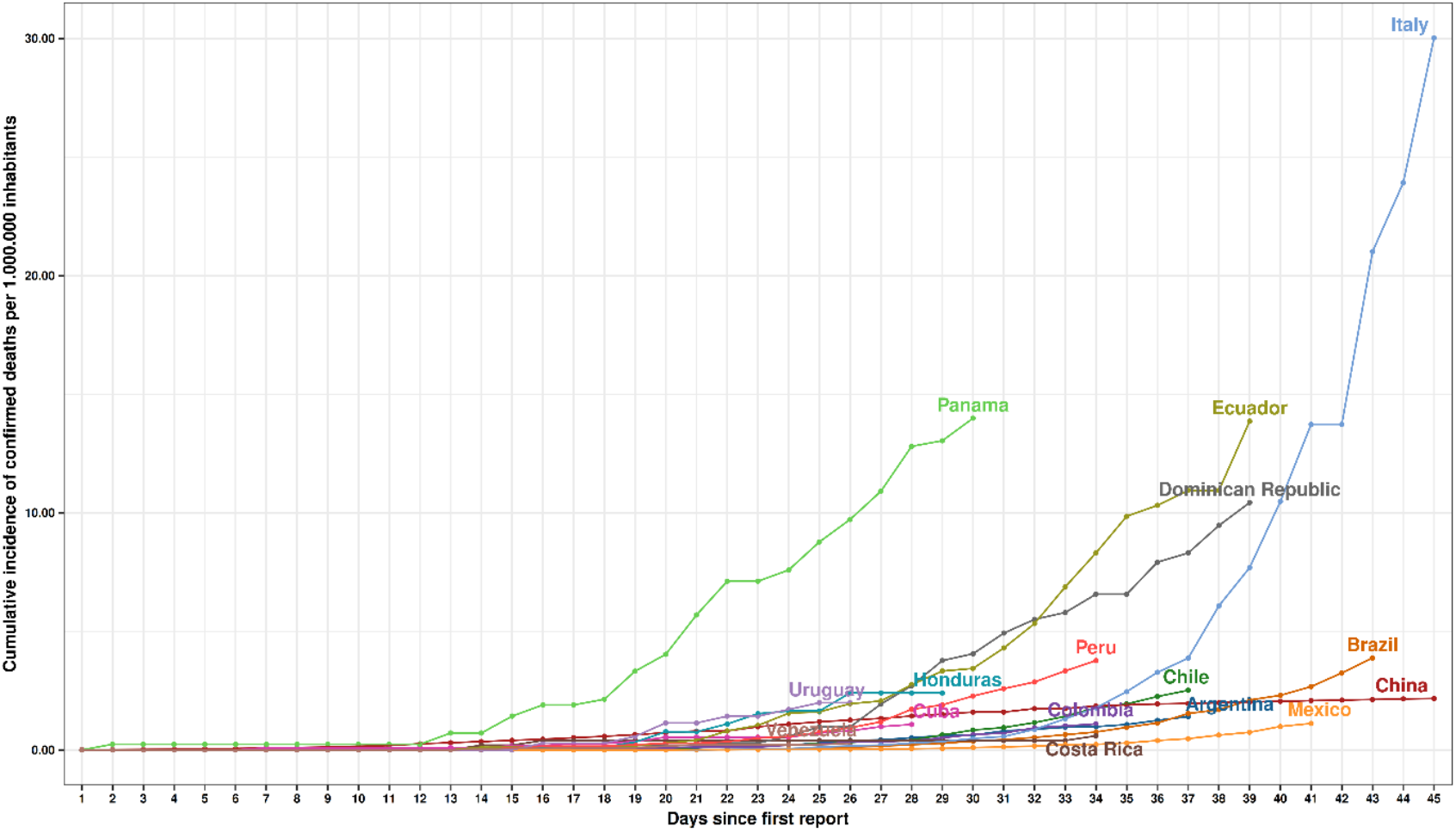
Cumulative incidence of confirmed deaths per million inhabitants since the first death reported in each country of Latin America and the Caribbean.

#### Case Fatality Rate (CFR%) Analysis

The fatality rate from COVID-19 in Italy reached 12.6% on the 68^th^ day after the first case report. During the same time, China had a CFR% a little over 4.03% at 77^th^ day after the initial report (Table 1).

In Latin-American the computed CFR% ranged from 4% to 7.5% during the initial stage of the pandemic infection. Trinidad and Tobago has the highest CFR% with 7.48%. Meanwhile, it was followed by countries like Bolivia (7.14%), Honduras (7.05%) and Ecuador (5.44%). Chile (0.87%), Uruguay (1.65%) and Costa Rica (0.60%) reported the lowest CFR% within the region (Table 1).

### Testing-Related Data Analysis

#### Tests per Country per Million Inhabitants

In terms of testing, by April 8^th^, Italy reported a total of 807,125 performed tests. In addition, when it was corrected by population level, it demonstrated 13,349 tests performed per million inhabitants (TMI). In Latin America, the country with the highest number of tests performed was Chile with 60,391, followed by Brazil with 54,824 (Table 1), Peru with 39,599 and Colombia with 33,575 (Figure 7). Nevertheless, concerning the number of tests performed by population, Panama and Chile reported the highest testing rates per million people in the region (>2,500), followed by Uruguay and Peru with more than 1,200 TMI each. In contrast, Brazil, Mexico and Ecuador, countries that were the first in reporting COVID-19 cases in the region, could not reach more than 900 TMI.

**Figure 7.**
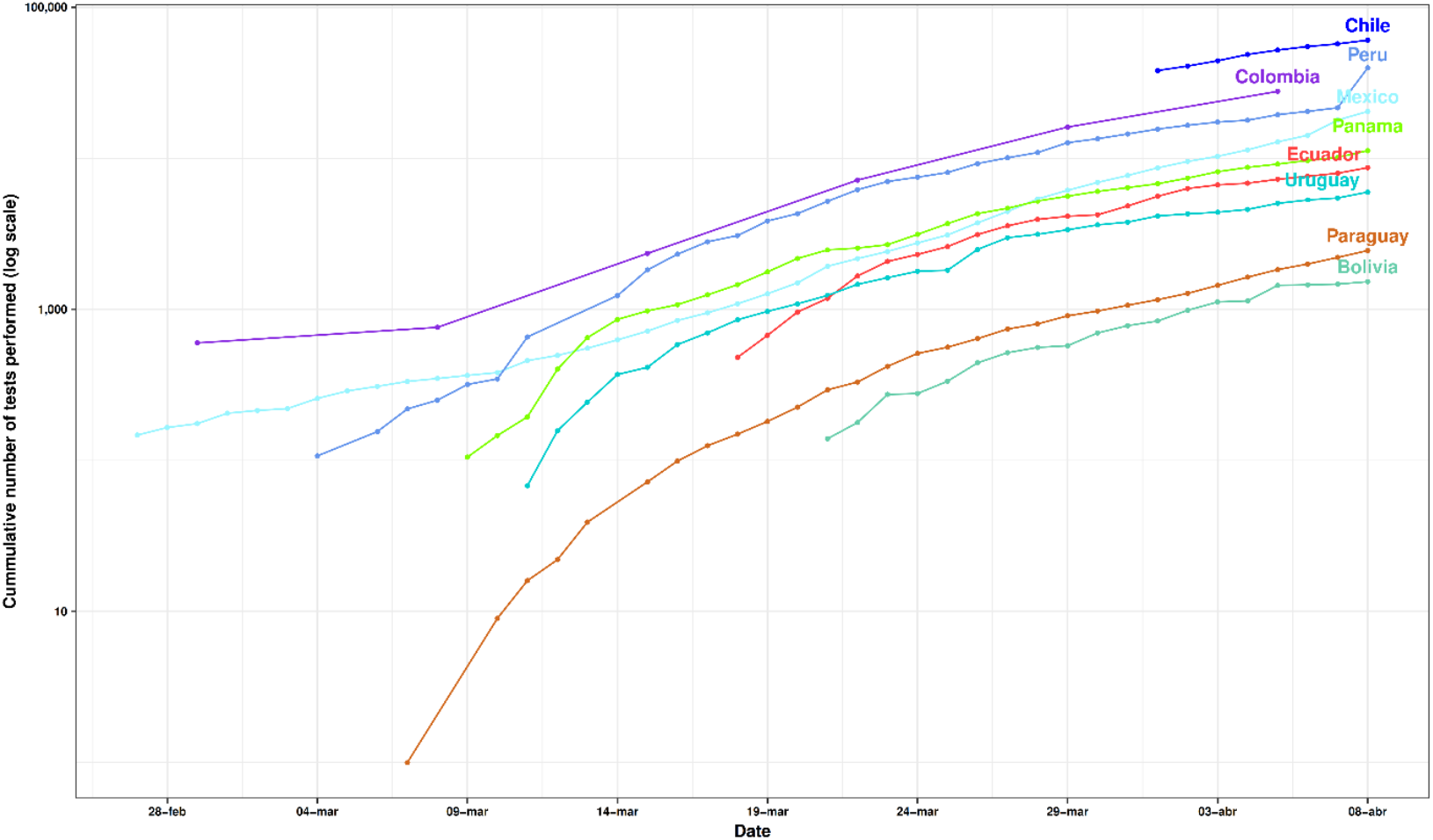
COVID-19 Tests performed in Latin-American countries. Source: Our World in data, 2020.

#### Daily Tests per Million People

On January 31^st^, Italy reported its first case of coronavirus, after 68 days, on April 8^th^, this country had reached its highest number of daily tests with at least 196 tests performed in a single day per one million inhabitants.

In Latin America, some countries kept the number of daily testing close to 100 test per day per one million people. For instance, Panama performed 94 daily test per one million people, followed by Chile and Uruguay, countries that have reached at least 90 daily test per one million people. On the other hand, some countries like the Dominican Republic, Bolivia, Ecuador and Brazil did not accomplish to reach at least 25 daily test per one million people (Table 1).

#### Positive Test Rate (PTR %) Analysis

The ratio between the number of tests and number of tests which resulted positive was analysed for Italy and Latin-American countries on April 8^th^. Italy reported a ratio of 17.27% of confirmed cases from the total tests performed (PTR%). In Latin America, some countries with highly effective testing strategies such as Chile and Uruguay have reported a positive test ratio of 9.18% and 7.38%, respectively (Table 1). On the other hand, countries with lower testing capabilities have reported higher positive testing rates. For instance, Dominican Republic reported the highest positive rate, up to April the 8^th^, with at least 46.4% of the total number of tests being positive, followed by Bolivia (35.5%), Ecuador (30.9%) and Brazil (29.5%) (Table 1).

### SARS-CoV-2 Genomes and Amino Acid Replacements in Latin America

Scientific communities from 52 countries have sequenced 4,347 SARS-CoV-2 genomes and submitted to GISAID between December 2019 and April 2020. Additionally, the CoV-GLUE Project has identified 2,334 amino acid replacements, 4 insertions, and 26 deletions. The most frequent amino acid replacements worldwide were D614G (S glycoprotein), P323L (nsp12), L84S (ORF8), Q57H (ORF3a), R203K (N phosphoprotein), G204R (N phosphoprotein), L37F (nsp6), T85I (nsp2), G251V (ORF3a), and Y541C (nsp13) (Figure 8A).

**Figure 8.**
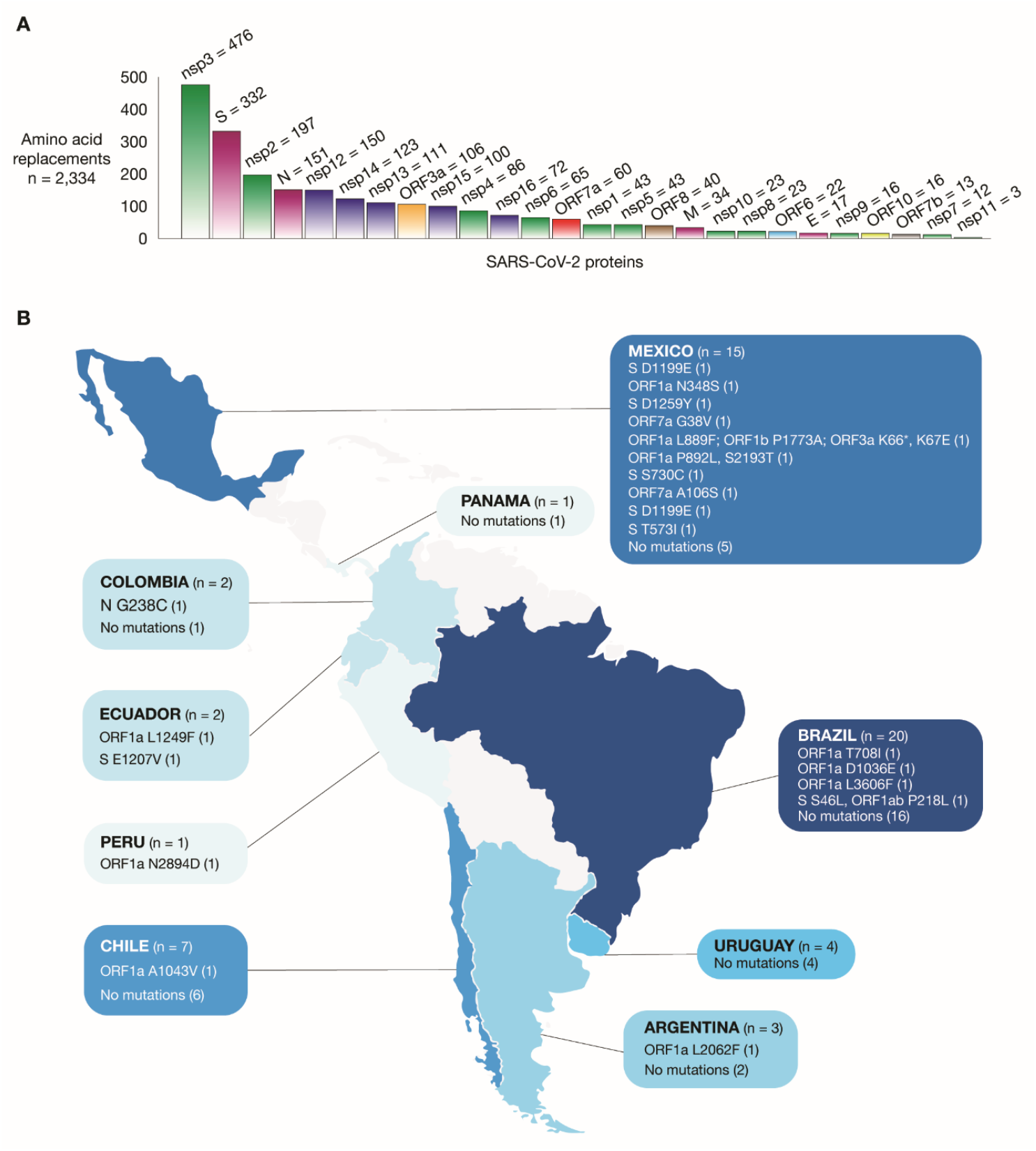
SARS-CoV-2 genomes in Latin America. A) SARS-CoV-2 proteins with the highest number of amino acid replacements. B) Amount of SARS-CoV-2 genomes and location of amino acid replacements per Latin American country.

Regarding Latin America, 55 SARS-CoV-2 genomes have been sequenced and submitted to GISAID. Brazil analysed 20 genomes finding the ORF1a T708I, ORF1a D1036, ORF1a L3606Fand other amino acid replacements shown in (Figure 8B). Mexico analyzed 15 genomes finding 10 amino acid replacements listed in (Figure 8B); Chile analyzed 7 genomes finding only 1 amino acid replacement; Uruguay analyzed 4 genomes without mutations; Argentina analyzed 3 genomes finding only 1 amino acid replacement; Ecuador analyzed 2 genomes finding 2 amino acid replacements; Colombia analyzed 2 genomes finding only 1 amino acid replacement; Peru analyzed 1 genome finding only 1 amino acid replacement; and, Panama analyzed 1 genome without mutations.

## Discussion

The Pan-American Health Organization and United Nations announced that Latin American and Caribbean countries had a slight time advantage on preparation for the coronavirus pandemic arrival [21, 22] due to the delay in seeing cases arising compared to other locations. LAC countries have some of the highest urbanization rates in the world, often with unequal access to public services in urban settings and high mortality rates for transmittable diseases [23, 24]. In some countries, out-of-pocket expenditure for healthcare services places a substantial restriction on access, debilitating viral control efforts [25, 26]. Furthermore, the region has experienced a wave of political and economic changes during the previous years and with the arrival of COVID-19, increased pressure is placed on already overstretched healthcare services. This combination of inequality, lack of governability, and debilitated services have already resulted in tragic scenarios during the COVID-19 pandemic in some LAC countries.

Our analysis depicts Panama as the most affected country in the region when a population correction is applied, not only due to the increase in positive cases (CCC, CICC) but also due to the number of official deaths reported (CCD, CICD). Panama in comparison with Italy, one of the most heavily affected countries in the world showed a dramatic increase in the number of cases, an exponential growth that was reached 18 days earlier than Italy. We speculate that as Panama has high aerial and maritime international traffic, these factors that might have played an important role importing asymptomatic cases earlier than its first report [27, 28].

Other countries have case-related data (CCC, CICC, DCMI) increasing at a faster rate than Italy. Chile, for instance, one of the richest countries in the region had a marked increase in the number of confirmed cases during the third week of the local out-brake. This situation could be explained due to the Chilean effective testing strategies [29, 30]. For instance, Chile performed 88 molecular test per million people every day with one of the lowest positive test ratios in the region (9.18%). Chile is also one of the countries with the lowest COVID-19 related deaths in the region (CCID, CCD and CFR) [31]. Uruguay had initially increased in a similar trend like Panama or Ecuador, nevertheless, as the days passed and more test arrived, this country showed cumulative rates similar to those seen in Chile [32].

Another fact about other LAC countries compared with Chile is that not enough tests are being performed, thus a sub-registration of COVID-19 is evident. Countries such as the Dominican Republic, Bolivia and Ecuador have limited testing capabilities [33], a fact that is supported by the low number of TMI, DTMI and the high PTR% in relationship with the number of tests performed.

Estimation of the real number of cases of the novel coronavirus (SARS-CoV2) infections is critical for understanding the frequency of presentation of COVID-19 and the overall pandemic potential of this infection [34]. A recent report from China suggested that at least 86% of all infections were not identified, while the transmission rate of undocumented cases was estimated to be at least 55% of the total of reported infections [34].

In the region, Ecuador has been highlighted due to its COVID-19 pandemic management and the rapid geographic spread of the novel coronavirus [35]. This equatorial country received worldwide attention due to the excessive number of undocumented deaths upon the arrival of the SARS-CoV-2 virus within the region [35]. According to the official figures published by the National Emergency Management Committee Ecuador reported in April 16^th^, at least 10,939 deaths were officially recorded during the first 45 days of the arrival of the virus in Ecuador (March to April 15^th^), a number 202% higher than the 3,622 deaths recorded during the first two months of 2020 [35]. These number is largely dependent on the number of cases reported in the province of Guayas, the province with the highest impact of the arrival of COVID-19 in the country. Although the government has not officially stated that the excessive numbers of deaths are due to COVID-19, a significant number of these deaths are likely COVID-19 related, especially when other violent causes of deaths (traffic accidents and homicides) have been reduced due to the partial cuff implemented in the country during the first week of March [36–38].

Brazil, the most populated country in the region, had the highest number of cases reported in the region, nevertheless, when the population-size was corrected a reduction in CICC, CICD, TMI, DTMI and DCMI values were observed. Brazil has been struggling with a lack of strong political leadership to minimize the effects of COVID-19 within the communities which undoubtedly has contributed to the high daily confirmed cases per million inhabitants.

Concerning the positive test rate percentage within the region, the 10% threshold suggested by the World Health Organization is an indicator that helps us determine the diagnosis efforts performed for each country seems to be too high to accomplish for several countries. For instance, Dominican Republic reported the highest positive rate in April the 8^th^ with at least 46.4% of the total number of tests being positive, followed by Bolivia (35.5%), Ecuador (30.9%) and Brazil (29.5%). When we compare these figures with other countries, we can observe that Ecuador, one of the most affected in the region has almost 120% more positive cases per every test performed than Italy, 1,500% more than South Korea and almost 7,000% more than Vietnam[30].

It is important to empathize that the figures reported rely on national government statistics, nevertheless, is critical to understand that testing capabilities are widely reduced in some countries in the region. Without adequate mechanisms of efficiently testing as much population as possible, contact tracing strategies, and effective isolation of suspected cases becomes a real challenge[29].

The analysis of these trends is an interim insight into the current situation in the region. The best action that countries can take is implementing social distancing measures and reducing travel, so individuals are less likely to come into contact with each other, therefore reducing the rate of transmission and alleviating pressure on health systems.

## Limitations

During the performance of this study, a disparity of the reports obtained from the different freely accessible published databases was evidenced regarding the reporting dates of the first case, confirmed cases and registered deaths values. For this reason, to maintain uniformity of the data, Johns Hopkins University database was used for graphs and tables.

Regarding the rates used, it’s evident that CFR% and PTR% are highly variable during the pandemic, therefore they don’t reflect correctly the impact of deaths and tests carried out in a country. Furthermore, the recovered case data was not used because, until the end of the study, any country reported more than a thousand cases, and it is suggested that these variables and the adjusting for outcome delay in CFR estimates should be analysed in subsequent studies.

It is important to understand that there is lack of reliable diagnostic tests (RT-PCR) data in the region. On top of that, some countries do not report the results of private providers, thus the analysis is often performed with the available data.

Another critical limitation is the lack of information for most of the countries when distinguishing between tests that were collected versus those which were processed.

Understanding the role of good testing strategies is important for the correct understanding of the health crisis at the individual and collective level. The lack of installed capabilities in some countries can produce a false sensation of fewer cases, while there are more cases out there, cases that cannot be identified [34].

This study only used confirmed cases of COVID-19, and undoubtedly there will be a large proportion of individuals who have died without testing conducted or are in remote communities which may go unreported.

## Conclusions

The experience and trends from Italy showed that the COVID-19 pandemic is progressing rapidly in LAC Countries. Thus, hospitals should prepare themselves for an increase in the number of patients with COVID-19 requiring healthcare, and in particular intensive care units where patients may require ventilation. Most LAC counties have implemented different public health measures to decrease the pressure on healthcare since the third week of March to mitigate the impact of the pandemic, such as travel restriction and social distancing. Finally, there is a wide variety of scenarios established between LAC countries, however, in this study, there was a lack of testing for a number of countries. This is an issue which needs addressing, and requires strong political will and resource to be able to provide the best care for patients and their families.

## Data Availability

All the data is publicly available through the servers that were described within the methodology section.

https://coronavirus.jhu.edu/map.html

## Abbreviations

CCC: Cumulative Confirmed Cases
CCD: Cumulative Confirmed Deaths
CICC: Cumulative Incidence of Confirmed Cases
CICD: Cumulative Incidence of Confirmed Deaths
CFR%: Case Fatality Rate
COVID-19: Novel Coronavirus Disease
CSSE: Center for Systems Science and Engineering
DCMI: Daily Cases per Million Inhabitants
DTMI: Daily Tests per Million Inhabitants
GISAID: Global Initiative on Sharing All Influenza Data
LAC: Latin America and Caribbean
MERS: Middle-East Respiratory Syndrome
SARS: Severe Acute Respiratory Syndrome
TMI: Tests per Million Inhabitants
PTR%: Positive Test Rate

## Credit Author Statement

KSR, LGB, EOP conceived the idea of the Editorial. The rest of the authors reviewed and improved the first and second draft. All authors approved the final version.

## Author Contributions

KSR, LGB and EOP were responsible for the original conceptualization of the project and they are fully responsible for every aspect of the current manuscript. They were fully involved in data acquisition and analysis.

JG and FSG were responsible for part of the data acquisition and they also contributed with the drafting of the manuscript.

RFN was fully responsible for generating the figures and part of the statistical analysis.

ALC was responsible for the genomic analysis within the region and the conceptualization and creation of figure eight.

AL critically reviewed and edited the manuscript, as well as offered important insights that strengthened the manuscript significantly.

## Ethical Approval

According to the local guidelines and good clinical practice, all secondary publicly available anonymized data does not require IRB approval.

## Acknowledgements

According to GISAID requirement, we included a supplementary Table acknowledging to all contributions and laboratories that are part of such an institution [16, 17]

## References

[1] Hays JN. Epidemics and Pandemics: Their Impacts on Human History. ABC-CLIO; 2005.

[2] Petrosillo N, Viceconte G, Ergonul O, Ippolito G, Petersen E. COVID-19, SARS and MERS: are they closely related? Clin Microbiol Infect Off Publ Eur Soc Clin Microbiol Infect Dis 2020. https://doi.org/10.1016/j.cmi.2020.03.026.

[3] Ortiz-Prado E, Simbaña-Rivera K, Gomez-Barreno L, Rubio-Neira M, Guaman LP, Kyriakidis N, et al. Clinical, Molecular and Epidemiological Characterization of the SARS-CoV2 Virus and the Coronavirus Disease 2019 (COVID-19): A Comprehensive Literature Review 2020. https://doi.org/10.20944/preprints202004.0283.v1.

[4] Zhu N, Zhang D, Wang W, Li X, Yang B, Song J, et al. A Novel Coronavirus from Patients with Pneumonia in China, 2019. N Engl J Med 2020. https://doi.org/10.1056/nejmoa2001017.

[5] Zhou P, Yang X-L, Wang X-G, Hu B, Zhang L, Zhang W, et al. A pneumonia outbreak associated with a new coronavirus of probable bat origin. Nature 2020. https://doi.org/10.1038/s41586-020-2012-7.

[6] WHO. Novel Coronavirus. Nov Coronavirus – China Dis Outbreak News Update 2020. http://www.who.int/csr/don/12-january-2020-novel-coronavirus-china/en/ (accessed April 9, 2020).

[7] Taylor DB. A Timeline of the Coronavirus Pandemic. N Y Times 2020.

[8] Coronavirus confirmed as pandemic. BBC News 2020.

[9] COVID-19 Map - Johns Hopkins Coronavirus Resource Center n.d. https://coronavirus.jhu.edu/map.html (accessed April 14, 2020).

[10] Gilbert M, Pullano G, Pinotti F, Valdano E, Poletto C, Boëlle P-Y, et al. Preparedness and vulnerability of African countries against importations of COVID-19: a modelling study. The Lancet 2020;395:871–877.

[11] McKibbin WJ, Fernando R. The global macroeconomic impacts of COVID-19: Seven scenarios 2020.

[12] Rodriguez-Morales AJ, Gallego V, Escalera-Antezana JP, Méndez CA, Zambrano LI, Franco-Paredes C, et al. COVID-19 in Latin America: The implications of the first confirmed case in Brazil. Travel Med Infect Dis 2020.

[13] WHO. Novel Coronavirus (2019-nCoV) situation reports 2020. https://www.who.int/emergencies/diseases/novel-coronavirus-2019/situation-reports (accessed April 8, 2020).

[14] Dong E, Du H, Gardner L. An interactive web-based dashboard to track COVID-19 in real time. Lancet Infect Dis 2020. https://doi.org/10.1016/S1473-3099(20)30120-1.

[15] Ritche H. Coronavirus Source Data. Our World Data 2020. https://ourworldindata.org/coronavirus-source-data (accessed April 22, 2020).

[16] Elbe S, Buckland-Merrett G. Data, disease and diplomacy: GISAID’s innovative contribution to global health. Glob Chall 2017. https://doi.org/10.1002/gch2.1018.

[17] Shu Y, McCauley J. GISAID: Global initiative on sharing all influenza data – from vision to reality. Eurosurveillance 2017. https://doi.org/10.2807/1560-7917.ES.2017.22.13.30494.

[18] CoV-GLUE. Amino acid analysis for the SARS-CoV-2 pandemic 2020. http://covglue.cvr.gla.ac.uk/#/home (accessed April 10, 2020).

[19] Bank W. Combining the Quantitative and Qualitative Approaches to Poverty Measurement and Analysis: the Practice and the Potential. Washington, DC: World Bank; 1997.

[20] United Nations. Population, Environment and Development: The Concise Report. United Nations Publications; 2020.

[21] Mitchell C. OPS/OMS | El tiempo para desacelerar la propagación de la COVID-19 se está acortando en las Américas, los países deben actuar ahora. Pan Am Health Organ World Health Organ 2020. https://www.paho.org/hq/index.php?option=com_content&view=article&id=15762:el-tiempo-para-desacelerar-la-propagacion-de-la-covid-19-se-esta-acortando-en-las-americas-los-paises-deben-actuar-ahora&catid=740:pressreleases&lang=es&Itemid=1926 (accessed April 13, 2020).

[22] WHO. Coronavirus disease 2019 (COVID-19). Situation Report – 83. WHO; 2020.

[23] Colón-González FJ, Peres CA, São Bernardo CS, Hunter PR, Lake IR. After the epidemic: Zika virus projections for Latin America and the Caribbean. PLoS Negl Trop Dis 2017;11:e0006007.

[24] Shepard DS, Undurraga EA, Halasa YA, Stanaway JD. The global economic burden of dengue: a systematic analysis. Lancet Infect Dis 2016;16:935–941.

[25] Atun R, De Andrade LOM, Almeida G, Cotlear D, Dmytraczenko T, Frenz P, et al. Health-system reform and universal health coverage in Latin America. The Lancet 2015;385:1230–1247.

[26] Ortiz-Prado E, Ponce J, Cornejo-Leon F, Stewart-Ibarra AM, Trujillo RH, Espín E, et al. Analysis of health and drug access associated with the purchasing power of the ecuadorian population. Glob J Health Sci Internet 2017;9.

[27] Redacción. La insólita medida de Panamá contra el coronavirus: separar las salidas de hombres y mujeres. BBC News Mundo 2020.

[28] Sabonge R, Sánchez R. El Canal de Panamá en la economía de América Latina y el Caribe 2009.

[29] Torres I, Sacoto F. Localising an asset-based COVID-19 response in Ecuador. The Lancet 2020.

[30] Total COVID-19 tests conducted vs. Confirmed cases. Our World Data n.d. https://ourworldindata.org/grapher/covid-19-total-confirmed-cases-vs-total-tests-conducted (accessed April 22, 2020).

[31] Paúl F. Cómo Chile ha logrado mantener a raya el covid-19 (y cuál puede ser su talón de Aquiles). BBC News Mundo 2020.

[32] Redacción. Uruguay considera ser uno de los países que “mejor está tratando” el covid- 19 | El Comercio. El Comer 2020.

[33] Navarro J-C, Arrivillaga-Henríquez J, Salazar-Loor J, Rodriguez-Morales AJ. COVID- 19 and dengue, co-epidemics in Ecuador and other countries in Latin America: Pushing strained health care systems over the edge. Travel Med Infect Dis 2020. https://doi.org/10.1016/j.tmaid.2020.101656.

[34] Li R, Pei S, Chen B, Song Y, Zhang T, Yang W, et al. Substantial undocumented infection facilitates the rapid dissemination of novel coronavirus (SARS-CoV2). Science 2020.

[35] Ecuador’s Response to the Coronavirus «Economics «Cambridge Core Blog n.d. https://www.cambridge.org/core/blog/2020/04/21/ecuadors-response-to-the-coronavirus-pandemic/ (accessed April 22, 2020).

[36] COE Nacional. Informes de Situación e Infografias – COVID 19 – desde el 29 de Febrero del 2020 – Servicio Nacional de Gestión de Riesgos y Emergencias 2020. https://www.gestionderiesgos.gob.ec/informes-de-situacion-covid-19-desde-el-13-de-marzo-del-2020/ (accessed April 18, 2020).

[37] Guayaquil reporta más casos de COVID-19 que 7 países de la región. El Universo 2020. https://www.eluniverso.com/guayaquil/2020/04/02/nota/7802199/guayaquil-reporta-mas-casos-covid-19-que-7-paises-region (accessed April 22, 2020).

[38] Coronavirus | COVID-19 | Desproporcionado índice de muertes abre interrogante sobre el alcance de la COVID-19 en Ecuador | RPP Noticias n.d. https://rpp.pe/mundo/actualidad/coronavirus-covid-19-desproporcionado-indice-de-muertes-abre-interrogante-sobre-el-alcance-de-la-covid-19-en-ecuador-noticia-1259602 (accessed April 22, 2020).

